# State of health of the gut microbiome assessed through a quantitative metabolomic-based marker panel

**DOI:** 10.1101/2025.02.14.24319815

**Authors:** Orlando DeLeon, Mary Frith, Ashley M Sidebottom, Jason Koval, Karen Lolans, Hugo D Ceccato, Candace M Cham, Na Fei, Elizabeth Wall, Sonia Kupfer, Arial Sims, Joseph F Pierre, Eugene B Chang

## Abstract

Defining the “health” of the gut microbiome or microbial organ has been elusive and controversial. This in turn makes defining gut dysbiosis (imbalances in gut microbiota) an enigma particularly in determining what, how, and when to intervene to restore eubiosis. Taxonomical profiles of gut microbiota have typically been used but are unreliable because of highly variable interindividual taxa and arbitrary functional assignments generalized to microbial groups. We therefore developed a functional metabolomic marker panel, selected through a supervised, machine-learning approach, using a healthy adult discovery cohort. The panel’s performance and utility were evaluated individually and together using a composite Z score through; (1) two validation cohorts of self-described healthy adults, 2) a large, external inflammatory bowel disease cohort, and 3) a patient with functional bowel disease to direct dietary intervention. The findings of this study provide proof of concept that quantitative functional profiles of the gut microbiome can be useful in assessing the health of the gut microbiome and for clinical application.

## Introduction

The gut microbiome is a vital organ of the body, essential for fostering and maintaining human health^1–3^, and when imbalanced (termed dysbiosis) can cause or contribute to many diseases^4–8^. However, unlike other bodily vital organs that have diagnostics to assess their function, defining the state of health of the gut microbiota through 16S rRNA amplicon sequencing [which only capture microbiome membership and relative abundance^9,10^] or by predictive functional assignments [based on taxonomical classification] have not been clinically useful^11–15^. Alternative genomic technologies that indirectly measure function include shotgun metagenomics and metatranscriptomics, wherein all bacterial genes or transcripts are sequenced. However, these technologies are time-consuming to analyze, expensive, and bacterial mRNA are highly labile making them impractical for clinical application. These limitations have prompted the microbiome field to increasingly adopt metabolomics to directly measure functional output of the microbiome^16^. While gut microbial community composition can be highly variable across individuals and populations, its function is likely conserved due to universal processes that the gut microbiome must provide (e.g., digestion of complex carbohydrates, conversion of bile acids, fermentation pathways, etc.). Thus, this functional readout can yield the closest representation of phenotype^16,17^.

Herein we describe the development of a stool-based metabolomics tool to evaluate the health status of the gut microbiome. To adopt an inclusive definition of health, 82 healthy adult subjects defined as lacking debilitating or medically unmanageable disease provided stool from which we analyzed 300+ targeted metabolites known to be produced or modified by the gut microbiota. From the 82 subjects, “Unhealthy” datasets (shadow sets) were randomly generated that were ±1, ±2, and ±3 standard deviations (SD) for each metabolite. We then selected metabolite features based on a supervised neural nets approach, wherein a set of 20 features were randomly selected from major molecular classes representative of microbiome function, lowest population variation, and measured for their performance. To generate a single score representative of the total microbiome function, we computed the composite Z-score for these 20 metabolites, which we term “the score”. We then evaluated its usefulness by 3 methods. First, we examined two cohorts; one cohort containing 14 healthy subjects with multiple timepoint sampling, and a second cohort containing 21 healthy subjects with single sampling to verify that the range of the score was not affected in independent healthy cohorts. Second, we utilized a large, external metabolomics dataset of healthy, Ulcerative Colitis, and Crohn’s disease subjects to determine the ability of our panel to correctly discriminate healthy vs unhealthy subjects. Last, we used the score as a management tool to monitor a single patient with functional bowel disease during a 45-week course of dietary interventions.

## Results

### Machine Learning (ML)-based selection of microbiome metabolites and development of normal reference ranges to monitor gut microbiota health

The stool metabolome is a direct measurement of microbiome function, and thus, is closest to phenotype. However, a challenge in the development of metabolomics-based diagnostics has been the identification of features indicative of microbial dysfunction, but consistent within individuals and the population. We measured the abundance of 300+ metabolites known to be modified or produced by the microbiome in the cohort of 82 individuals lacking debilitating disease and self-describing as ‘healthy’ (Table 1). While the chemical space of the microbiome is diverse^18,19^, these metabolites span many distinct chemical classes including amino acids, bile acids, simple and complex carbohydrates, fatty acids, indols and tryptophan derivatives, organic and aromatics and TCA intermediates.

**Table 1.** Demographic and clinical characteristics of N=82 subjects.

|  | *N* | Percent (%) |
| --- | --- | --- |
| **Gender** |  |  |
| Male | 33 | 40.24% |
| Female | 49 | 59.76% |
| **Age** |  |  |
| Average | 38 (+/-13) |  |
| 18-30 | 31 | 37.80% |
| 31-40 | 24 | 29.27% |
| 41-50 | 11 | 13.41% |
| 51-60 | 8 | 9.76% |
| 61-80 | 8 | 9.76% |
| **BMI** |  |  |
| Average | 24.2 (+/- 3.46) |  |
| <18.5 (underweight) | 2 | 2.44% |
| 18.5-24.9 (healthy) | 49 | 59.76% |
| 25.0-29.9 (overweight) | 22 | 26.83% |
| >29.9 (obese) | 7 | 8.54% |
| unlisted | 2 | 2.44% |
| **Race** |  |  |
| Asian | 13 | 15.85% |
| Black or African American | 2 | 2.44% |
| Hispanic or Latinx | 6 | 7.32% |
| Multi- or Bi-Racial | 5 | 6.10% |
| White or Caucasian | 53 | 64.63% |
| Other | 3 | 3.66% |

To capture the intra-individual and intrapopulation variation of microbial metabolites, 3 stool samples were provided by each individual within a 7 day period, irrespective of prandial status. We calculated the standard deviation (SD) for each metabolite within each individual, or the intraindividual variation, and plotted the values in Log2 scale (Fig. 1, A). The population level variation for each metabolite (population SD) was calculated by repeat random sampling of each individual with replacement (nperm=100). We found that variation of a metabolite within an individual was a property specific to the metabolite irrespective of its chemical class and consistent between individuals (i.e. the degree of variation for a metabolite was similar between subjects, but the variation could be consistently high or low). This correlated to the population-level variation of a given metabolite, with features of high interindividual variability also having high intrapopulation variability (ie the range of values for a metabolite could be very large or low, but specific to each metabolite). To identify metabolites useful for evaluating the functional output of the microbiome, we applied a supervised machine learning approach with several constraints (Fig. 1, B). First, we understand these data to represent the range of a given metabolite in a healthy population and thus reasoned dysfunctional microbiomes or “unhealthy” microbiomes would be outside this range, departing from the population average the greater the dysfunction. To train an ML algorithm to distinguish health vs unhealth with this defition, we created an unhealthy cohort by randomly altering markers from randomly selected individuals from the healthy cohort to be outside the range of the normal healthy reference ranges (+/−1, 2, or 3 population-level SDs of the healthy population ranges), which we term our “unhealthy shadow population” dataset. By doing this, the ML algorithm could be trained to identify deviations from a healthy population as “not healthy” the farther from the population center value, and allows us test and validate our model. This also allows us to identify markers that work well in combination to identify eubiosis and dysbiosis in a general population. Second, we understood that adaptation to a clinical setting requires that the number of metabolites be limited and finite (to reduce cost and turnaround time) but still capture the breadth of functions provided by the microbiome to affect health. We applied a second constraint of 1-5 selected features within a given molecular class, limiting the number of metabolite features to 20 based on the following considerations. We evaluated the ability of features to discriminate health from unhealth by iteratively testing sets of 20 features (nperm=1000) and comparing their feature importance scores (decrease in Area Under the Curve, AUC) using a machine learning, neural nets methodology (dividing our datasets into 70:30, test:train, with replacement). Last, we performed a power calculation to determine the number of subjects needed to confirm the population average and range of each metabolite (at 80% confidence) and excluded features that required more than 300 subjects. (Inspection of these final 20 metabolites confirmed a standard normal distribution or a similar chi-square distribution, allowing for the calculation of ranges and confidence intervals in Supplemental Figure 1.) This led to a final panel of 20, top-scoring metabolites that span multiple molecular classes, perform well to discriminate functional status of the microbiome, and can be validated with a reasonable number of subjects (Fig. 1, C).

**Figure 1.**
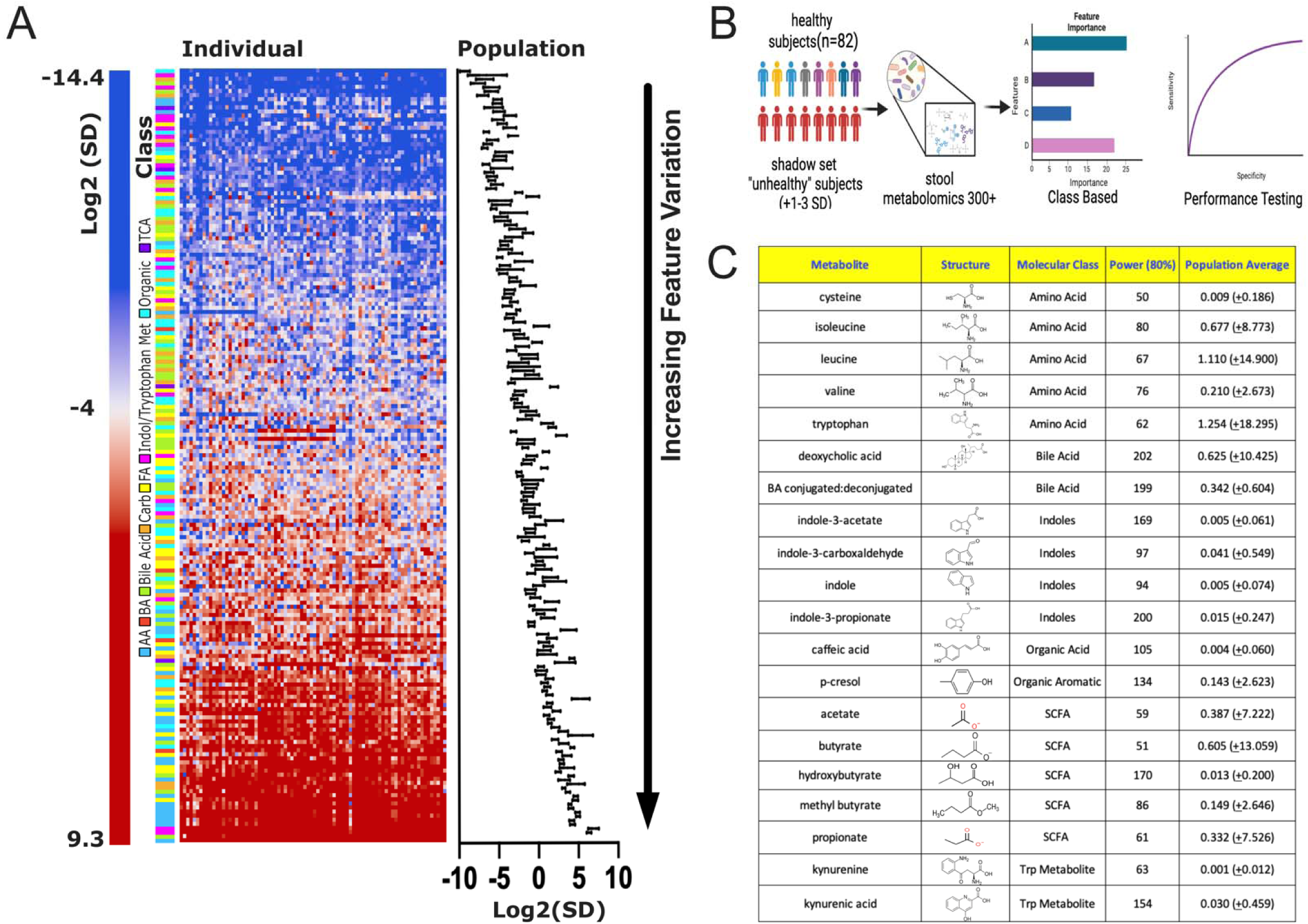
ML-based selection of microbiome-based metabolites to measure health. 300+ stool metabolites known to be produced or modified by the microbiome were analyzed by mass spectroscopy. 100 signals that fell below the limit of detection for >50% of the population were removed. Intraindividual variation was calculated for each metabolite and compared to its population variation (Log2(SD)), demonstrating that metabolites with high variation in an individual correlated to the population variation (**A**). Schematic for 20 metabolite panel selection (**B**). An “unhealthy dataset” was informatically generated by randomly selecting a sample from an individual and introducing +/− 1, 2, of 3 population SDs to the feature value, which we term the “shadow set”. We then used machine learning to identify 20 features/metabolites able to discriminate between the healthy and unhealthy datasets applying a selection constraint of 1-4 metabolites from each molecular class for each performance testing. The final panel of 20 markers with their population and chemical characteristics (**C**).

Our choice to adopt a more inclusive definition of health was to generate a tool capable of monitoring microbiome health in a general population. However, we wished to determine whether these reference ranges were significantly altered if we adopted a stricter definition of health. We calculated the reference ranges of each metabolite from n=62 subjects that had no prior history of GI or metabolic disease and found no difference in either the means or distributions (T-test and F-test, respectively, p>0.05 for each metabolite) (Supplemental Table 1). Thus we conclude that for these metabolites, inclusion of the entire dataset was warranted.

### Evaluation of a minimal (20) metabolite ML classifier score for defining gut microbiome health

To evaluate the ability of our metabolomics-based ML classifier, we performed repeat test-and-train of the data (nperm=1000, 70:30, test:train) using the normal ranges established for each of the 20 features. We plotted the false positive rate (1-Specificity) against the true positive rate (Sensitivity) for the categorical classes of “healthy” and “unhealthy” (Fig. 2, A). We found the AUC=0.924 with contributions by each metabolite to the performance of the classifier ranging from 0.4 (indole-3-carboxaldehyde) to 0.08 (deoxycholic acid, DCA) (Fig. 2, B). This suggests that even the least important feature, DCA, contributed significantly to the classification capacity. Thus we arrived at the selection of a minimal 20 feature panel with high performance and representative of multiple metabolomic functions.

**Figure 2.**
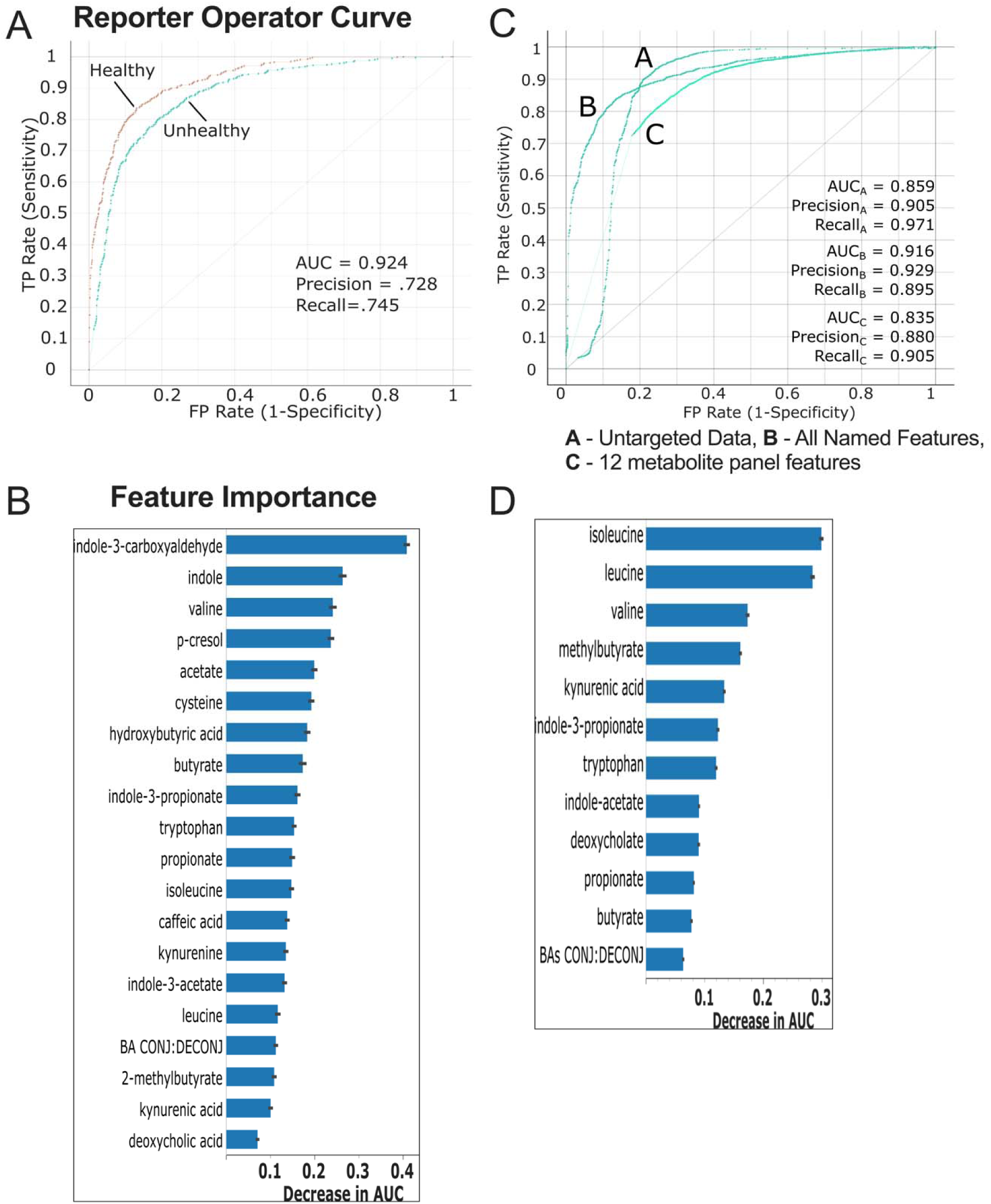
Validation of a minimal (20) metabolite ML classifier for microbiome health. Reporter operator curves (ROCs) for the classification of healthy vs unhealthy (**A**). Repeat test and train (70:30) of the data was performed to test the ML classifier using the 20 features. FP Rate (Specificity) and TP Rate (Sensitivity) are plotted for the identification of each class, with an AUC= 0.924. Feature Importance of the 20 metabolite signals are shown as decrease in AUC (**B**). An independent metabolite dataset consisting of healthy, Crohn’s disease and ulcerative colitis subjects was used to generate and test (permutated, randomized test and train) ML-based classifiers using the untargeted metabolomics feature data (curve A), all the named metabolite features (curve B), or a minimal 12-member panel based on the shared features with our in-house panel (curve C) (**panel C**). The 12-member panel performed well with an AUC_C_=0.835 compared to AUC_A_=0.859 for the untargeted data and AUC_B_=0.916 using all the named features, demonstrating the capacity of our panel to discriminate healthy vs unhealthy subjects. Feature importance shown as decrease in AUC is shown for the 12-member panel of shared features (**D**).

Next, we assessed the ability of the metabolites to distinguish healthy vs unhealthy using a large, publicly available metabolomics dataset of healthy and IBD subjects (Metabolomic Workbench ‘ST000923’)^20,21^. This study included several hundred subjects classified as healthy (n=134), or diagnosed with Crohn’s disease (CD, n=266) or ulcerative colitis (UC, n=146). Due to differences in the examined metabolites institutions, we identified 12 metabolite features that were common between the two studies including isoleucine, leucine, valine, tryptophan, deoxycholate, indole-3-acetate, butyrate, propionate, methylbutyrate, kynurenic acid, indole-3-propionate, and the conjugated:deconjugated BA ratio. We tested the reduced 12-member panel on this independent cohort and found that the classifier could distinguish between healthy and unhealthy individual (UC or CD) (AUC=0.835) compared to using a classifier using all the named metabolites (AUC=0.916) or the untargeted feature data (AUC=0.859) (repeat test:train, 80:20, nperm=100) (Fig. 2, B). These features ranged in importance between 0.3-0.06 (decrease in AUC). Together, these data demonstrate that the selection of markers could distinguish between gut eubiosis and dysbiosis.

Selection of informative markers may be skewed by gender-differences^22^. Therefore, we also explored how the selected markers were affected by gender. Using Principal Component Analysis (PCA) using the 20 metabolite features, we found no significant difference between men and women (Fig. 3, A, PERMANOVA>0.05). Within the 20-metabolite panel, no differences were found in abundance between men and women, with the exception of deoxycholic acid (Fig. 3, B, 1-way ANOVA) and no difference in the intraindividual variation between men and women (Fig. 3, C, Student’s T-test). However, metabolite features not included in our 20-member panel were found to be skewed by gender, including 4-hydroxyphenyl acetate, hyodeoxycholic acid, isodeoxycholic acid, dihydrocaffeic acid, and muricholic acid (Supplemental Fig. 2). Therefore, while gender-differences exist in the overall metabolome of our training cohort, our selected panel markers are not sensitive to those differences. While our goal was to generate a general and broad tool, future panels specific to males and females may be useful to evaluate the gender-specific conditions.

**Figure 3.**
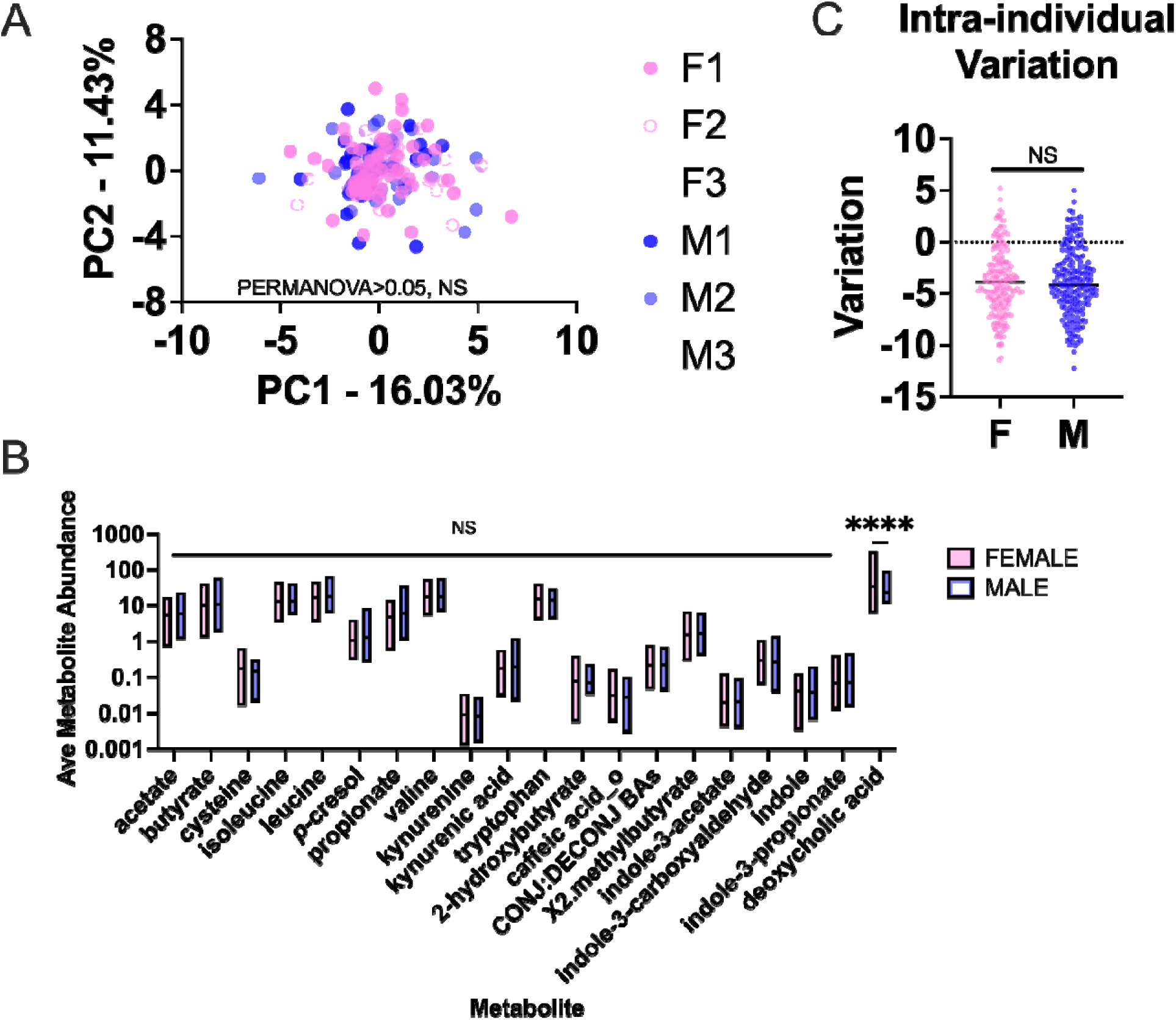
Gender-specific differences in metabolite panel. Principle component analysis shows no statistical difference (PERMANOVA>0.05) between men (blue) and women (pink) using the 20 metabolite featur s or between the first, second and third samples provided (F1vM1, F2vsM2, F3vsM3) (**A**). Abundances of each metabolite was not different between men and women for the 20 metabolite features selected (Tukey’s post-hoc, <0.05), with the exception of deoxycholic acid (Tukey’s post-hoc, *p<0.05) (**B**). Intraindividual variation (calculated for each individual form the 3 samples provided) was not significantly different between genders for each metabolite (Wilcoxon matched-pairs signed rank test, p=5042) (**C**).

### Evaluation of a composite score representative of microbiota health in independent, healthy cohorts

The metabolomic panel can provide a wealth of information, but to make it useful as a screening tool and interpretable for patients and medical providers, we derived a composite gut microbiota health score by calculating the average for the Z-scores of the 20 features from the median for each metabolite of the panel. We calculated the scores for the 82 inclusively healthy training cohort subjects vs the unhealthy shadow set and found that the distribution of healthy subjects fell below 20, with the vast majority (95%) below 10, suggesting a natural cutoff for “normal” (Fig. 4, A). Analysis of the frequency distribution of scores in healthy subjects showed 50% with scores of ≤3 and 95% ≤5.5 (Fig. 3, B). In the artificially generated unhealthy shadow subjects created from randomly inserting and altering deviated values of metabolites used to train our ML algorithm, 50% had scores of ≤14, 75% ≤25, and 95% ≤ 38, suggesting ranges of scores for full function are within ≤5, moderate function 5-14, ≥14 as moderately dysfunctional, and ≥ 25 as severe dysfunction.

**Figure 4.**
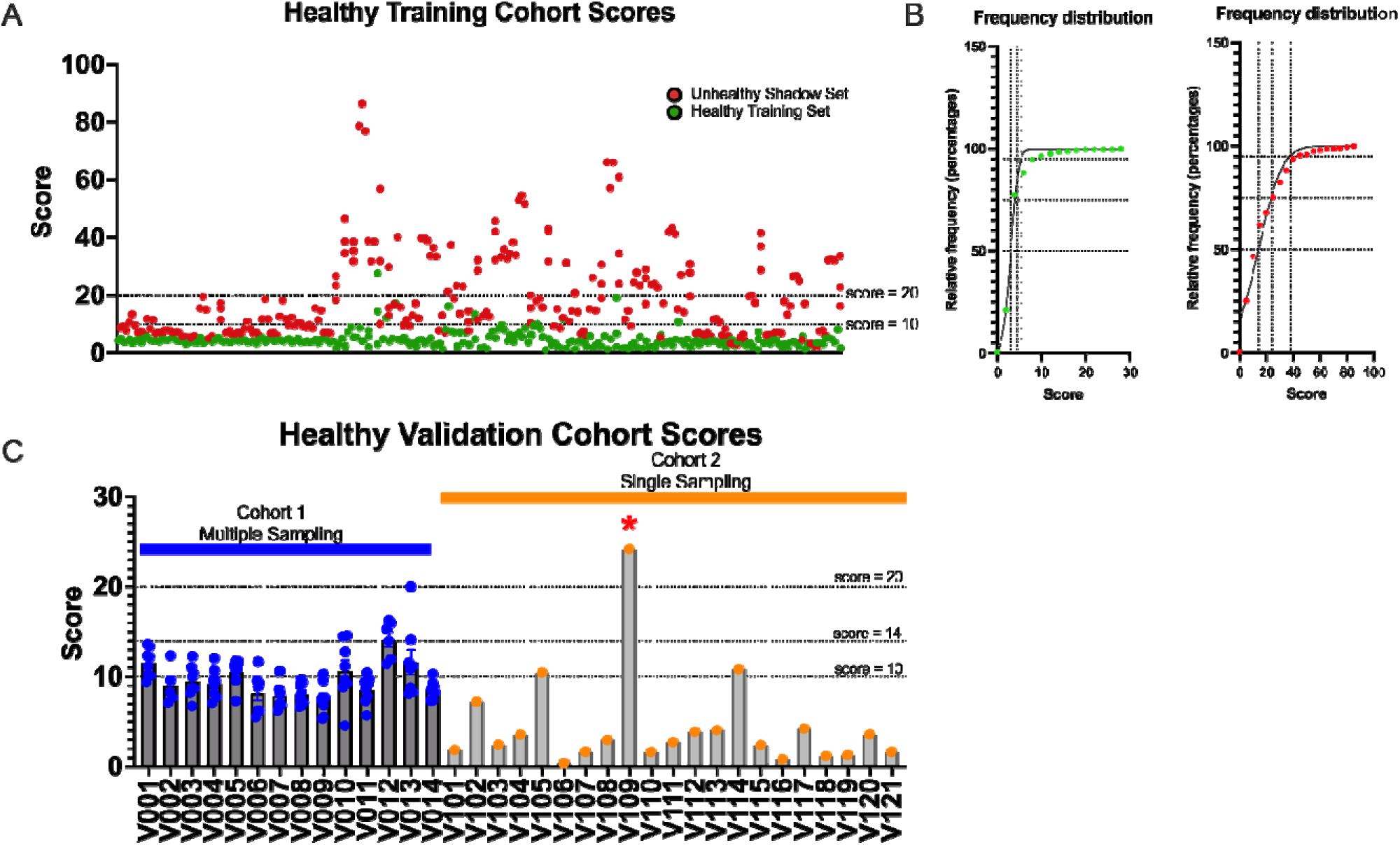
Evaluation of a composite score representative of microbiota functional health in independent, healthy cohorts. While the ML classifier is valuable in identifying healthy vs unhealthy subjects, it requires significant expertise to employ which can limit clinical use. We created an easy to use metabolomics score based on the ML-selected model features based using a composite, unsigned Z-score, or “score”. Metabolomic composite scores for our healthy and unhealthy training cohort series show 20 as a cutoff for unhealthy and >10 as a potential indicator of microbiome functional deviation (**A**). Gaussian distribution of healthy and unhealthy training sets to determine score cutoff (**B).** Relative frequency percentages of 50, 75 and 95% are shown with the corresponding scores for healthy and unhealthy population sets. Examination of independent cohorts with multiple sampling (blue) and single sampling (orange). 2-independent, self-reported healthy cohorts were used to examine the stability of the healthy vs unhealthy score cutoffs (**C**). Patient V109 presented with >20 score, indicating a deviation from a healthy gut metabolome and was found to have hypercholesteremia and hypertension.

We next validated the utility of the score by examining other healthy subjects. First, we wished to examine whether natural intrapersonal variation affected the score distribution and our cutoffs of 10 and 20 were viable. We examined an independent cohort 14 self-reported healthy human subjects with repeat sampling (n=8 samples) over the course of 28 days and 21 self-reported healthy human subjects with single sampling. We found that the distribution of scores generally below 14 for each individual (Fig. 4, C), suggesting that the score cutoffs were robust. 3 individuals (V010, V012, and V013) had multiple samples within the intermediate range of possible dysfunction (14<20). One subject, V019, presented with a score 25. While this patient initially self-reported as healthy, but was later recategorized because of a high BMI (33), hypercholesteremia, hypertension, and the use of statins and ACE inhibitors. We also examined the scores of the IBD-cohort analyzed in Fig. 2 using 12 of the 20 metabolite features and found that CD and UC patients had significantly higher scores compared to healthy individuals (Supplemental Fig. 3, 1-way ANOVA, p<0.05*) Together, these data suggest natural ranges from healthy, possible dysfunction, and severe dysfunction can be identified. Furthermore, it highlights that we can identify deviations of normal microbiome function with single samples.

### Use of the metabolomics panel and score as a patient management tool, a case study

Practical use of a metabolomics tool is paramount for its adoption in a clinical setting. Therefore, we set up a use case for the metabolomics panel and overall score to determine how well they performed in management and treatment of a patient with a functional gastrointestinal disorder (FGID), “Patient Q”. FGID is a diagnosis of exclusion based on a constellation of recurring symptoms not specific to other disease presentations and without a known underlying molecular etiology^23,24^. Typical symptoms of FGID include chronic abdominal discomfort, bloating, constipation, diarrhea and/or stool irregularity. Patient Q presented with hallmark symptoms of a FGID (e.g., irregular bowel movements, gas, loss of continence, and food intolerance and abdominal discomfort) (Fig. 4, A). Onset of his symptoms occurred 10 years ago when 12-months into an 18 month-long course of doxycycline, the patient developed symptoms that led to chronic disturbances in daily activity, increased anxiety and depression, and an overall loss of quality of life. Exhaustive GI evaluations at several medical centers were unrevealing. Considering onset of symptoms coincided with extended use of antibiotics, it was suspected that gut dysbiosis had developed and that Patient Q’s food intolerances were a function of improper microbial output. Patient Q was referred to the University of Chicago for microbiome assessment and a dietary intervention designed to reconstitute his microbiome. We used the 20-metabolite panel to guide a 45-week dietary intervention program designed to reintroduce food types with self-reporting of GI-symptoms and thrice weekly monitoring of the microbiome by 16S rDNA sequencing and metabolomics.

Dietary intervention utilized the reintroduction phase of the FODMAP elimination diet ^25,26^ after careful analysis of Patient Q’s habitual diet consisting mostly of animal proteins and foods with low fermentable carbohydrates, patient-identified trigger foods, medications, and dietary supplements. Incidents of constipation, diarrhea, gas, bloating, pain or discomfort were reported (Fig. 5, A). In brief, the diet introduced one fermentable carbohydrate group or food-type per week with stepwise increases in serving size. Specifically, Day 1 allowed for one serving of a fermentable carbohydrate, Day 2 allowed for two servings, and Days 3-5 allowed for three servings. On days 6 and 7 Patient Q had the option to continue eating the recently introduced foods or return to his habitual diet. If a particular food or fermentable carbohydrate provoked unpleasant GI symptoms, the food was immediately eliminated from the intervention. He reported significantly reduced but intermittent discomfort, gas, and incidences of food intolerance as well as reduced duration of discomfort from several days to several hours. Over the 45-week intervention, Patient Q reported a significant reduction in his GI symptoms that allowed for a reduction in medications used to manage his FGID (50% reduction of polyethylene glycol 3350, 67% reduction of gabapentin, and elimination of amitriptyline). While we do not have direct evidence that we completely restored his microbiome which resulted in symptomatic improvement, our tool helped was critical in informing and guiding management towards gut eubiosis.

**Figure 5.**
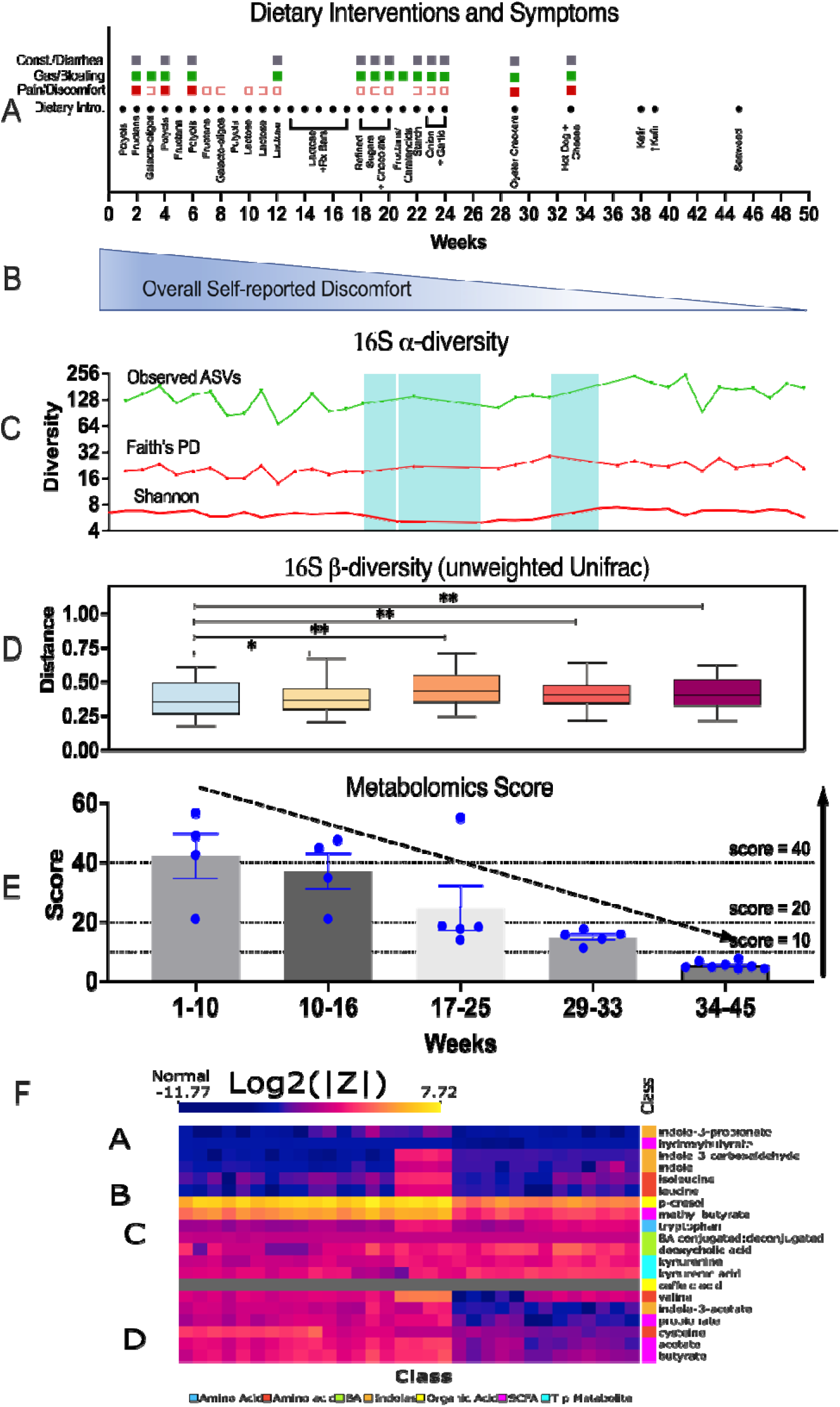
Utilization of the metabolomics panel score as a patient management tool, a use case. A white, male in thier 70s with functional bowel disorder was monitored by 16S rRNA and stool metabolomics during a 45 week series of dietary interventions. Microbiome metrics demonstrate the utility of our metabolomics panel relative to 16S-based metrics. 50 week course of dietary interventions marked by patient reported incidences of GI pain/discomfort, gas/bloating, and constipation/diarrhea (**A**). Self-reported discomfort over the cours of the treatment window. (**B**) 16S-based alpha diversity metrics for observed amplicon sequence variants (ASVs), Faith’s phylogenetic diversity, and Shannon’s diversity. Blue highlighted regions represent periods of time during which alpha diversity differed from baseline (Mann-Whitney, p<0.05). (**C**) show no significant changes with the exception of weeks 18-26 and 32-34 (Mann-Whitney, P>0.05). 16S-based beta diversity changes (unweighted UniFrac) distances show significant differences relative to weeks 1-10 (PERMANOVA, *P<0.05) but not specific trajectory. Metabolomics composite score demonstrates a reduction over time, suggesting a return to “normal” microbiome function (**E**). Normalization of specific panel metabolites relative to the population average (**F**). |Z-score| for each metabolite was calculated relative to the population average for Patient Q over time (Log2 scale). Hierarchical clustering of metabolites revealed 4 groups. A-remaining close to the population average, B-and D-markedly returned to the population average, and C-not changed and not near the population average.

Assessments of his microbiome by 16S revealed no significant change to his alpha diversity by the number of ASVs, Faith’s phylogenetic diversity, or Shannon’s metric, suggesting no change occurred (either increase or decrease) in the membership of his gut microbiome (Fig. 5, C). Assessment of his β−diversity demonstrated a change from baseline in the unweighted UniFrac distances (Fig. 5, D). However, no pattern or trajectory was detected in the changes, suggesting that while the proportions of his microbiota shifted during the course of the dietary intervention, they did not do so in a concerted, consistent treatment-dependent manner. Initial assessment of his metabolome by our panel revealed a high score of 42.22 (+/−7.62) in the first ten weeks of monitoring, suggesting his microbiome functional output was abnormal (Fig. 5, E). Over the course of the dietary intervention, we found a steady, detectable decrease in his score resulting in a final score of 5.354 (+/− 0.44). We then identified which metabolites were most abnormal and how they changed over the course of the dietary intervention (Fig. 5, F). We visualized the Log2 of the absolute value of the Z-score relative to the population average and performed hierarchical clustering of the metabolite features. We found 4 clusters that remained normal over time (A), were markedly distinct from the population but returned to normal levels (B and D), or remained distinctly different from the population (C). Importantly, this allowed us to identify normalizations of *p*-cresol, methyl-butyrate, valine, indole-3-acetate, propionate, cysteine, acetate, and butyrate, suggesting a correction of fermentation and inflammatory processes. Overall, reductions in his score provided a readily interpretable (return to normal) metric by which the patient could assess his own improvement in microbiome functional output. In this case, this feedback provide strong motivation to continue with dietary intervention. For physicians, changes in key functional subsystems of the gut microbiome can be followed to assess response to the therapy.

## Discussion

For most vital organs of the body, multiple modalities that can determine states of health or disease are available. However, clinically useful diagnostic tools for the gut microbial organ (also called the gut microbiome) are lacking. This unsolved problem has encumbered both research investigations and the development of effective interventions for altered states of the gut microbiome (loosely defined as dysbiosis), the latter including probiotics, fecal microbiota transplantation, and other forms of therapeutics ^27–30^. Most commercial applications and research investigations of the gut microbiome have relied upon 16S rRNA amplicon profiles which provide genus level taxonomical information, but offer little insight into function. In addition, 16S rRNA amplicon profiles are highly variable among healthy individuals^31–33^, making is difficult to define health in quantifiable terms. Arguably, functional measures such metagenomics and metabolomics can be more informative because they provide insights of how microbiomes impact their host under conditions of health and disease. However, these tools have not been used clinically because of cost, required expertise for data reduction, difficulty in collecting accurate samples, turnaround time, and incomplete or unavailable annotation of metagenomic and metabolomic data sets. To address these issues, we developed a quantitative metabolomics panel of 20 markers, each representative of specific functional subsystems of the gut microbiome of healthy adults. An additional criteria for selection of the panel markers was inter-individual homogeneity whereby normal reference ranges could be established through an initial training cohort of 82 healthy adults (defined as absence of debilitating or unmanaged disease). This tool is comprised of a ML-based classifier as well as and easy-to-use score to evaluate the component and overall functional status of the gut microbial organ. We evaluated this tool in external datasets, independent cohorts of healthy individuals, and describe a specific use case in the management of a patient with FGID, Patient Q. We demonstrate that this tool and approach can be developed as a clinically useful tool to assess the state of health of the gut microbiome for patients with dysbiosis. Importantly, we have generated a framework to answer a much debated question in the microbiome field and developed a quantitative metric to define eubiosis and dysbiosis.

One of the major clinical challenges is patient adherence to treatment programs, particularly when treatment response metrics are not available. Therapies can be difficult or not-well tolerated and thus can cause patients to become discouraged. In our use case, our tool contributed to patient confidence in the treatment plan with easily interpretable and sensitive results compared with 16S-based microbiome technologies. Patient Q was able to quantify the level of improvement over time and qualify the functions in which he improved, often before symptomatic improvement. By contrast, changes to specific microbes were less directly interpretable and did not demonstrate obvious, distinct patterns of improvement.

We strove to make a panel for the general evaluation of microbiome function. Thus we adopted a broad, inclusive definition of health as “nondebilitating disease”. We show proof of concept that our panel and approach performs well to assess the state of gut microbiota health of a health Midwest population. In the future, this panel can be extended to develope disease-, population-, and demographic-specific diagnostics and classifiers. This includes the identification of novel metabolites through untargeted metabolomics that are more sensitive to specific diseases. We demonstrate that a limited panel of 12 metabolites generated to evaluate general microbiome function was capable of distinguishing healthy individuals from patients diagnosed with Crohn’s disease or ulcerative colitis (Fig. 2, C-D) highlighting our approach in defining unhealth as a deviation from a normal metabolite range using an independently conducted validation cohort. While CD and UC specific markers are likely to exist (with greater specificity and/or sensitivity), the discriminatory capacity of our panel demonstrates its general utility as an evaluation tool for health (if not necessarily suitable for diagnosis). Further studies examining how this panel discriminates between active vs inactive inflammatory disease states as well as other extra-intestinal disease states are required. Due to differences in methodologies, metabolites examined, and sample collection/preservation methods across institutions, publicly available data is limited. Thus, establishment of standardized methodology for microbiome metabolomics and complimentary computational methods for the correction of batch effects would greatly contribute to this effort and aid in further validation of our tool.

Last, a limitation of this study was its development from an adult (18-80), urban, American, Midwestern population and skewed towards a younger demographic of university students (aged 18-30 constitute 38.80%). Differences in the early life (0-5 years) or late life (>65) microbiomes may warrant the development of age-specific panels that can predict developmental outcomes or risk for decline. Differences in US regional populations and global populations reflect genetic, dietary, environmental, and lifestyle differences which affect microbiome composition and function. Likely, populations-specific panels will need to be developed that are sensitive and specific to the variability inherent to these populations. While development of these panels represent a significant research undertaking, it may also be an opportunity to learn which microbiota functions are variant or conserved between populations and thus describe universal, fundamental relationships between the human species at large and the gut microbiome.

## Data Availability

All data produced in the present study are available upon reasonable request to the authors

## Author Contributions

EC, ASidebottom, OD, NF, JP, JK, CC, and KL conceptualized and designed the study. OD, ASidebottom, HC analyzed the data. MF, OD, and EC wrote the manuscript. ASims and SK managed and treated Patient Q using the metabolic panel. All authors contributed to editing the manuscript. KL, HC, and JK collected and processed the samples for microbiome analysis. EC and EW managed the patient described in Figure 5, Patient Q.

## Methods

### Sample Collection

All human subjects in this study were consented and approved by the University of Chicago Institutional Review Board (IRB 22-0822). Stool samples were collected from all four datasets [healthy training cohort aka cohort of 82 healthy subjects (Fig. 3A), validation cohorts aka14 and 21 self-described healthy donors (Fig. 3C) and Patient Q (Figure 4)], using the EasySampler Stool Collection Kit (Alpco, cat#58-EZSAMPLER). Briefly, Patient Q collected stool samples thrice, weekly (Wed, Sat, & Sun) for 45 weeks over the course of a rigorous dietary intervention. Samples were stored in Patient Q’s personal freezer until transport to University of Chicago and stored at −80°C. Stool from the 14 and 21 self-described healthy donors were self-collected twice weekly for 4 weeks (8 samples total) or as single samples, respectively, with samples directly frozen after the time of collection. For the cohort of 82 subjects, three samples were collected within the span of 7 days. To allow for at home stool collection, each collection tube was modified by adding 3-mL of a 95% ethanol-based preservative. The preservative was spiked with two heavy molecules [0.49mM Caffeine-(trimethyl-d9) and 1.15mM sodium D3 acetate] to serve as internal analysis controls. Pilot studies demonstrated metabolite stabilization for 2 weeks at 22C and 37C (data not shown). The tubes were weighed before and after the addition of preservative to determine the exact stool mass submitted and allow for precise downstream metabolite quantification. All samples were thawed immediately prior to 16S rRNA or metabolomics.

### 16S rRNA sequencing

16S rRNA sequencing was performed at the University of Chicago Duchossois Family Institute in the Microbiome Metagenomics Core Facility (DFI MMF). DNA extraction from stool was performed using the QiaAmpPowerFecal Pro DNA (Qiagen, cat. # 51804) and the V4-V5 region of 16S rRNA-genes PCR amplified using barcoded dual-index primers (520FWD-AYTGGGYDTAAAGNG and 909REV-CCGTCAATTYHTTTRAGT). Illumina compatible libraries were generated using the Qiagen QIASeq 1-step amplicon kit (Qiagen, cat. #180419 and sequencing performed on an Illumina MiSeq platform using a 2×250 Paired-End reads, generating 5,000-10,000 reads per sample. Data analysis was performed on the QIIME2 platform^34^. Raw V4-V5 16S rRNA gene sequence data were demultiplexed and processed through the dada2 pipeline into Amplicon Sequence Variants (ASVs). Taxonomic assignments of ASVs up to the genus level were generated through a Naïve Bayes classifier trained on Silva database16S rDNA sequences (Silva v132) and were BLASTed against RefSeq for species-level identification with corresponding alignment statistics. Alpha diversity (Total ASVs, Shannon’s index, Chao’s metric, and Faith’s phylogenetic diversity) and beta diversity metrics (Bray-Curtis, weighted UniFrac, and unweighted UniFrac distances) and statistical significance were calculated in QIIME2 (Mann-Whitney and PERMANOVA, n=999).

### Metabolomics

Metabolomics was performed at the University of Chicago Duchossois Family Institute in the Host-Microbe Metabolomics Core Facility (DFI HMMF). The metabolome of stool was analyzed across four mass spectrometry platforms to capture quantitative and qualitative levels of gut-derived metabolites with varying physiochemical properties such as hydrophobicity, size, and charge. The DFI Host-Microbe Metabolomics Facility (DFI-HMMF) routinely studies intestinal and fecal material with the proposed methods and analysis pipelines. In brief, metabolites were extracted with organic solvent, dried down and resuspended for direct analyses or derivatization. Previously, all 305 compounds have been validated by the DFI-HMMF through retention time and fragmentation comparison to standards and available databases. Compounds were chosen based on known host-microbe mechanisms or the compound level in fecal material was shown to significantly vary across patient populations indicating a potential role in health. Gas chromatography-mass spectrometry (GC-MS) was used to detect 222 compounds following derivatization with pentafluorobenzyl bromide (PFBBr)^35^ and trimethylsilyl-methoxamine (TMS-MOX)^36,37^ in two separate reactions. SCFAs (acetate, butyrate, propionate), lactate, and succinate were quantitatively analyzed following PFB derivatization and detection by negative collision induced-gas chromatography-mass spectrometry ((−)CI-GC-MS, Agilent 8890). Another 48 PFB-derivatized compounds within the SCFA, branched chain fatty acid, amino acid, aromatic, hydroxylated fatty acid, organic acid, and indole compound subclasses were studied by normalized peak area. Positive ion electron impact-GC-MS ((+)EI-GC-MS, Agilent 7890B) was used to detect 169 molecules in the organic acid, carbohydrate, TCA intermediate, sterol, amino acid, indole and fatty acid subclasses following TMS-MOX derivatization. With the use of negative mode liquid chromatography-electrospray ionization-quadrupole time-of-flight-MS ((−)LC-ESI-QTOF-MS, Agilent 6546), 49 bile acids from the primary, secondary and glyco/tauro-conjugated subclasses were analyzed ^38^. In addition to retention time validation, the standard intact and fragment masses were routinely detected with differences < 5 ppm compared to calculated values. Positive mode LC-triple quadrupole-MS ((+)LC-ESI-QQQ-MS, Agilent 6547) were used to analyze 34 indole and tryptophan catabolites. Tryptophan is an essential amino acid that is ingested in the host diet. Conversion by microbes and the host result in biologically active compounds such as serotonin, kynurenine, and melatonin.

### Statistical and Machine Learning Methods

Metabolites were classified into molecular classes including bile acids, conjugated vs non conjugated bile acids, amino acids, tryptophan metabolites, fatty acids, tryptophan metabolites, indoles, and organic acids or aromatics. Downstream metabolomics values were corrected for batch effects using ‘*combat’* in R. All statistical analysis and data manipulation was performed in python (pandas, skit-learn v1.4, NumPy) with custom parsers, python-based data analysis platform Orange (v3) for the generation of heatmaps and secondary validation of the ML classifiers, and Prism v10 for Gaussian regression analyses and the calculation of the population means, median and standard deviations. Python ‘*dyplyr’* was used to randomly sample 1 of 3 sample per subject and introduce a random 1-3 population SD for the given metabolite to generate the “unhealthy” dataset or shadow set and categorically assigned as the target variable. Constraints for molecular classes (limited to 5 per class) were added for the random selection of 20 features which iteratively used in a neural nets model to calculate feature importance (ReLu activation, 100 hidden layers, L-BFGS-B solver, alpha=0.0001 regularization, replicable training, 200 iterations). Cumulative scores of feature importance for each metabolite over 1000 permutations were calculated to reduce the features and retested. The final 20 features were repeat test-train at a 70:30 split of the data. Overlapping features between our panel and the UC/Crohn’s dataset were first normalized using ‘*combat’* in R before use as a test set for the NeuralNet classifier (Metabolomic Workbench ‘ST000923’).

For all other plots and statistics (unless otherwise noted) were generated in Prism v10.

---

**Supplemental Figure 1.**
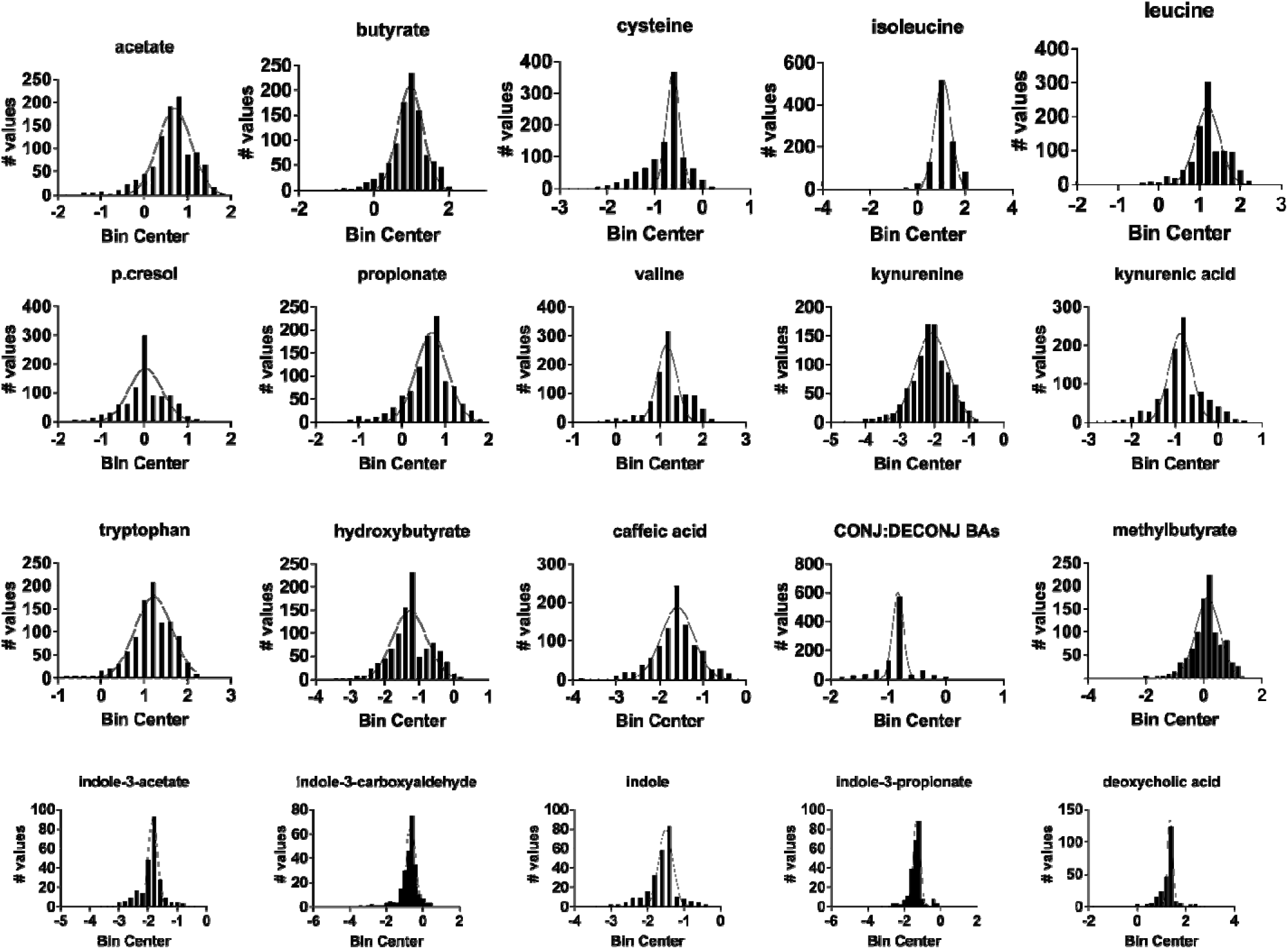
Population distributions of 20 panel metabolites in healthy subjects. Population distributions are shown for each of the 20 metabolite features for visual examination. All metabolites had a standard normal or a chi-square distribution.

**Supplemental Figure 2.**
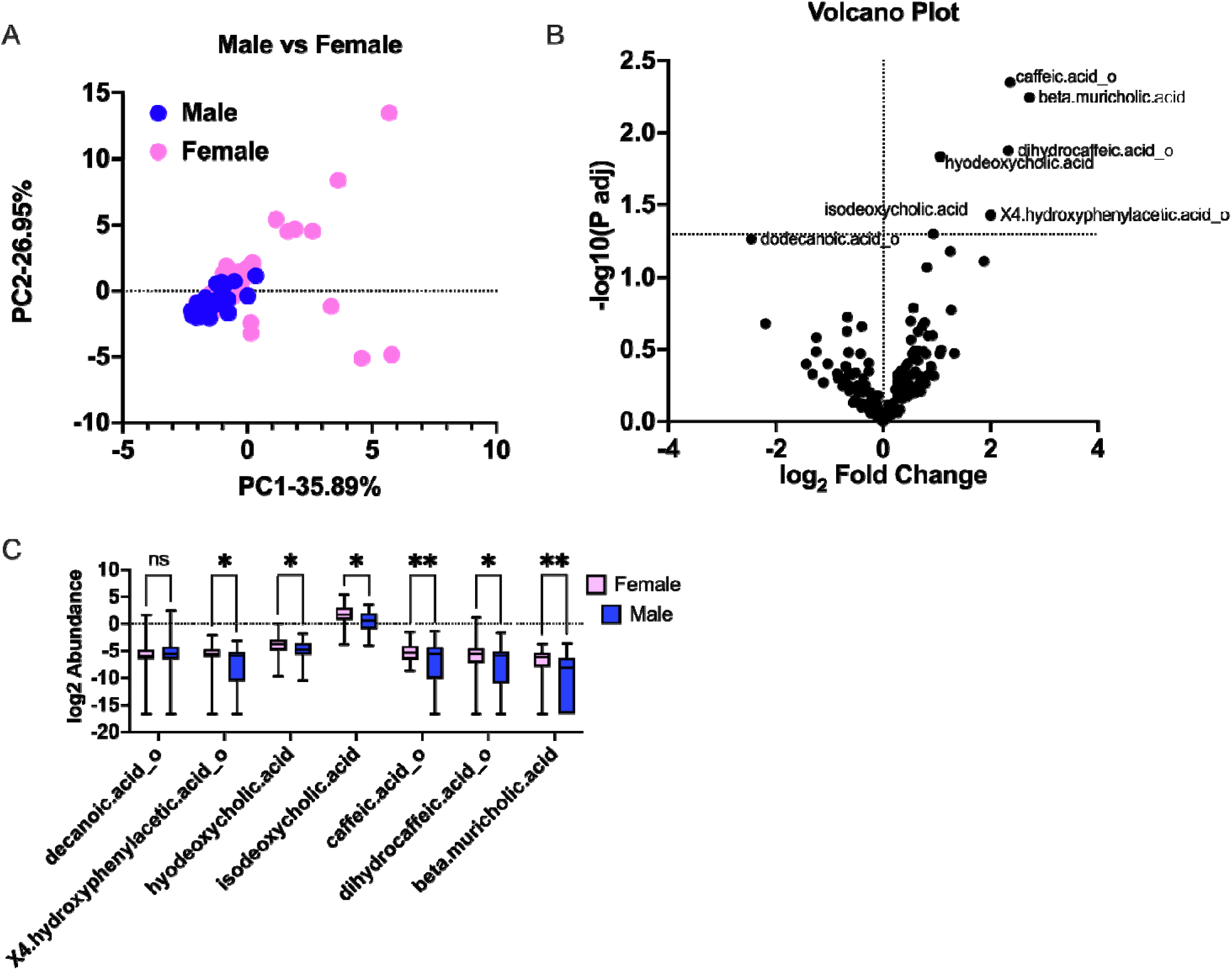
Identification of gender-specific metabolite signals. Principle component analysis (PCA) using all metabolite signals show no significant difference between genders, but a higher intragroup dispersion in females (**A**). Multiple metabolites were found to be significantly difference between men and women including muricholic acid, dihydrocaffeic acid, isodeoxycholic acid, hyodeoxycholic acid, and 4-hydroxyphenyl acetate show in a volcano plot (**B**) and box-and-whisker plot (**C**, 1-way ANOVA, p<0.05 for all pairwise comparisons).

**Supplemental Figure 3.**
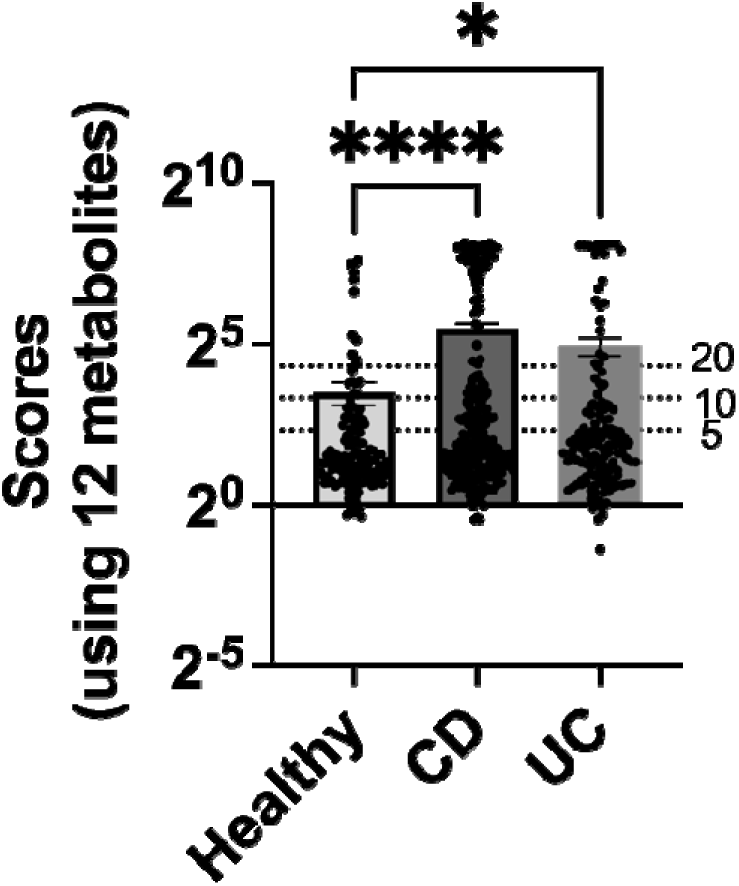
UC and CD patients have higher scores (based on 12 metabolites). Scores (log2 scale) were calculated based on the 12 metabolites shared between our panel and those within the IBD study (Metabolomics Workbench, ST00923). Scores were significantly higher for Crohn’s disease (CD) and ulcerative colitis (UC) patients than healthy controls (1-way ANOVA, *p<0.05 for healthy:CD and healthy:UC).

**Supplemental Table 1.** Comparison of metabolite ranges using inclusive (n=82) v exclusive (n=62) definitions of health. Population averages and standard deviations were calculated for the n-82 subject that met criteria for “non dbilitating disease” and for n=62 subjects that met strict criteria of health with no history or active GI or metabolic disease. T-stats, F-stats, and p-values were calculated to determine whether the stricter definiton of health affected the refernce range for metabolites used in this panel. T-statistics and two-tailed p-values were calculated to determine whether a statistical difference in the means exist. F-statistics and p-values were calculated to determine whether a statistical difference in variance exist.

| Metabolite | n=62 AVE | n=62 SD | n=82 AVE | n=82 SD | T-stat | p-value (two-tailed) | F-statistic | p-value |
| --- | --- | --- | --- | --- | --- | --- | --- | --- |
| cysteine | 0.011 | 0.176 | 0.009 | 0.186 | 0.027 | 0.647 | 1.123 | 0.638 |
| isoleucine | 0.602 | 7.956 | 0.677 | 8.773 | 0.155 | 0.877 | 1.216 | 0.425 |
| leucine | 1.009 | 13.635 | 1.110 | 14.900 | 0.160 | 0.873 | 1.194 | 0.470 |
| valine | 0.185 | 2.367 | 0.210 | 2.673 | 0.093 | 0.926 | 1.275 | 0.321 |
| tryptophan | 1.316 | 17.199 | 1.254 | 18.295 | 0.088 | 0.930 | 1.316 | 0.617 |
| deoxycholic acid | 0.784 | 10.620 | 0.625 | 10.425 | 0.291 | 0.772 | 1.038 | 0.869 |
| BA conj:deconj | 0.344 | 0.596 | 0.342 | 0.604 | 0.015 | 0.988 | 1.027 | 0.921 |
| indole-3-acetate | 0.006 | 0.053 | 0.005 | 0.061 | 0.018 | 0.986 | 1.310 | 0.271 |
| indole-3-carboxaldehyde | 0.050 | 0.554 | 0.041 | 0.549 | 0.071 | 0.943 | 1.019 | 0.927 |
| idole | 0.007 | 0.075 | 0.005 | 0.074 | 0.034 | 0.973 | 1.023 | 0.915 |
| indole-3-propionate | 0.018 | 0.232 | 0.015 | 0.247 | 0.031 | 0.976 | 1.134 | 0.610 |
| caffeic acid | 0.004 | 0.058 | 0.004 | 0.060 | 0.008 | 0.994 | 1.063 | 0.809 |
| p-cresol | 0.216 | 2.648 | 0.143 | 2.623 | 0.268 | 0.789 | 1.019 | 0.928 |
| acetate | 0.373 | 7.033 | 0.387 | 7.222 | 0.031 | 0.976 | 1.055 | 0.835 |
| butyrate | 0.769 | 13.258 | 0.605 | 13.059 | 0.268 | 0.790 | 1.031 | 0.891 |
| hydroxybutyrate | 0.017 | 0.183 | 0.013 | 0.200 | 0.048 | 0.790 | 1.191 | 0.476 |
| methyl mutyrate | 0.220 | 2.657 | 0.149 | 2.646 | 0.260 | 0.795 | 1.009 | 0.962 |
| propionate | 0.371 | 7.432 | 0.332 | 7.526 | 0.086 | 0.932 | 1.025 | 0.926 |
| kynurenine | 0.001 | 0.012 | 0.001 | 0.012 | 0.001 | 0.999 | 1.003 | 1.000 |
| kynurenic acid | 0.040 | 0.480 | 0.030 | 0.459 | 0.086 | 0.932 | 1.092 | 0.706 |

## References

1. Watson, A. R. et al. Metabolic independence drives gut microbial colonization and resilience in health and disease. Genome Biol. 24, 78 (2023).

2. Koppenol, E. et al. Fecal microbiota transplantation is associated with improved aspects of mental health of patients with recurrent Clostridioides difficile infections. J. Affect. Disord. Rep. 9, 100355 (2022).

3. Bull, M. J. & Plummer, N. T. Part 1: The Human Gut Microbiome in Health and Disease. Integr. Med. Encinitas Calif 13, 17–22 (2014).

4. Hou, K. et al. Microbiota in health and diseases. Signal Transduct. Target. Ther. 7, 135 (2022).

5. Wilkins, L. J., Monga, M. & Miller, A. W. Defining Dysbiosis for a Cluster of Chronic Diseases. Sci. Rep. 9, 12918 (2019).

6. Hrncir, T. Gut Microbiota Dysbiosis: Triggers, Consequences, Diagnostic and Therapeutic Options. Microorganisms 10, 578 (2022).

7. DeGruttola, A. K., Low, D., Mizoguchi, A. & Mizoguchi, E. Current Understanding of Dysbiosis in Disease in Human and Animal Models: *Inflamm*. Bowel Dis. 22, 1137–1150 (2016).

8. Winter, S. E. & Bäumler, A. J. Gut dysbiosis: Ecological causes and causative effects on human disease. Proc. Natl. Acad. Sci. 120, e2316579120 (2023).

9. Tikhonov, M., Leach, R. W. & Wingreen, N. S. Interpreting 16S metagenomic data without clustering to achieve sub-OTU resolution. ISME J. 9, 68–80 (2015).

10. Sun, Y. et al. A large-scale benchmark study of existing algorithms for taxonomy-independent microbial community analysis. Brief. Bioinform. 13, 107–121 (2012).

11. Jovel, J. et al. Characterization of the Gut Microbiome Using 16S or Shotgun Metagenomics. Front. Microbiol. 7, (2016).

12. Rajilić-Stojanović, M., Heilig, H. G. H. J., Tims, S., Zoetendal, E. G. & De Vos, W. M. Long-term monitoring of the human intestinal microbiota composition. Environ. Microbiol. 15, 1146–1159 (2013).

13. The Human Microbiome Project Consortium. Structure, function and diversity of the healthy human microbiome. Nature 486, 207–214 (2012).

14. Costello, E. K. et al. Bacterial Community Variation in Human Body Habitats Across Space and Time. Science 326, 1694–1697 (2009).

15. The Human Microbiome Project Consortium. A framework for human microbiome research. Nature 486, 215–221 (2012).

16. Bauermeister, A., Mannochio-Russo, H., Costa-Lotufo, L. V., Jarmusch, A. K. & Dorrestein, P. C. Mass spectrometry-based metabolomics in microbiome investigations. Nat. Rev. Microbiol. 20, 143–160 (2022).

17. Moya, A. & Ferrer, M. Functional Redundancy-Induced Stability of Gut Microbiota Subjected to Disturbance. Trends Microbiol. 24, 402–413 (2016).

18. Guthrie, L., Wolfson, S. & Kelly, L. The human gut chemical landscape predicts microbe-mediated biotransformation of foods and drugs. eLife 8, e42866 (2019).

19. Donia, M. S. & Fischbach, M. A. Small molecules from the human microbiota. Science 349, 1254766 (2015).

20. Metabolomics Workbench. PR000639. (2017) doi:10.21228/M82T15.

21. IBDMDB Investigators et al. Multi-omics of the gut microbial ecosystem in inflammatory bowel diseases. Nature 569, 655–662 (2019).

22. Costanzo, M. et al. Sex differences in the human metabolome. Biol. Sex Differ. 13, 30 (2022).

23. Thompson, W. G. Irritable bowel syndrome in general practice: prevalence, characteristics, and referral. Gut 46, 78–82 (2000).

24. Drossman, D. A. Functional Gastrointestinal Disorders: History, Pathophysiology, Clinical Features, and Rome IV. Gastroenterology 150, 1262–1279.e2 (2016).

25. Staudacher, H. M. & Whelan, K. The low FODMAP diet: recent advances in understanding its mechanisms and efficacy in IBS. Gut 66, 1517–1527 (2017).

26. Barrett, J. S. & Gibson, P. R. Fermentable oligosaccharides, disaccharides, monosaccharides and polyols (FODMAPs) and nonallergic food intolerance: FODMAPs or food chemicals? Ther. Adv. Gastroenterol. 5, 261–268 (2012).

27. Hoffmann, D. E. et al. The DTC microbiome testing industry needs more regulation. Science 383, 1176–1179 (2024).

28. Ghaffari, P., Shoaie, S. & Nielsen, L. K. Irritable bowel syndrome and microbiome; Switching from conventional diagnosis and therapies to personalized interventions. J. Transl. Med. 20, 173 (2022).

29. Acharjee, A., Singh, U., Choudhury, S. P. & Gkoutos, G. V. The diagnostic potential and barriers of microbiome based therapeutics. Diagnosis 9, 411–420 (2022).

30. Damhorst, G. L., Adelman, M. W., Woodworth, M. H. & Kraft, C. S. Current Capabilities of Gut Microbiome–Based Diagnostics and the Promise of Clinical Application. J. Infect. Dis. 223, S270–S275 (2021).

31. Turnbaugh, P. J. et al. Organismal, genetic, and transcriptional variation in the deeply sequenced gut microbiomes of identical twins. Proc. Natl. Acad. Sci. 107, 7503–7508 (2010).

32. Ursell, L. K. et al. The interpersonal and intrapersonal diversity of human-associated microbiota in key body sites. J. Allergy Clin. Immunol. 129, 1204–1208 (2012).

33. Neu, A. T., Allen, E. E. & Roy, K. Defining and quantifying the core microbiome: Challenges and prospects. Proc. Natl. Acad. Sci. 118, e2104429118 (2021).

34. Bolyen, E. et al. Reproducible, interactive, scalable and extensible microbiome data science using QIIME 2. Nat. Biotechnol. 37, 852–857 (2019).

35. Haak, B. W. et al. Impact of gut colonization with butyrate producing microbiota on respiratory viral infection following allo-HCT. Blood blood-2018-01-828996 (2018) doi:10.1182/blood-2018-01-828996.

36. Shan, J. et al. Integrated Serum and Fecal Metabolomics Study of Collagen-Induced Arthritis Rats and the Therapeutic Effects of the Zushima Tablet. Front. Pharmacol. 9, 891 (2018).

37. Fiehn, O. Metabolomics by Gas Chromatography–Mass Spectrometry: Combined Targeted and Untargeted Profiling. Curr. Protoc. Mol. Biol. 114, (2016).

38. Gómez, C., Stücheli, S., Kratschmar, D. V., Bouitbir, J. & Odermatt, A. Development and Validation of a Highly Sensitive LC-MS/MS Method for the Analysis of Bile Acids in Serum, Plasma, and Liver Tissue Samples. Metabolites 10, 282 (2020).

